# A Machine Learning Algorithm to Predict Hypoxic Respiratory Failure and risk of Acute Respiratory Distress Syndrome (ARDS) by Utilizing Features Derived from Electrocardiogram (ECG) and Routinely Clinical Data

**DOI:** 10.1101/2022.11.14.22282274

**Authors:** Curtis Earl Marshall, Saideep Narendrula, Jeffrey Wang, Joao Gabriel De Souza Vale, Hayoung Jeong, Preethi Krishnan, Phillip Yang, Annette Esper, Rishi Kamaleswaran

## Abstract

The recognition of Acute Respiratory Distress Syndrome (ARDS) may be delayed or missed entirely among critically ill patients. This study focuses on the development of a predictive algorithm for Hypoxic Respiratory Failure and associated risk of ARDS by utilizing routinely collected bedside monitoring. Specifically, the algorithm aims to predict onset over time. Uniquely, and favorable to robustness, the algorithm utilizes routinely collected, non-invasive cardiorespiratory waveform signals. This is a retrospective, Institutional-Review-Board-approved study of 2,078 patients at a tertiary hospital system. A modified Berlin criteria was used to identify 128 of the patients to have the condition during their encounter. A prediction horizon of 6 to 36 hours was defined for model training and evaluation. Xtreme Gradient Boosting algorithm was evaluated against signal processing and statistical features derived from the waveform and clinical data. Waveform-derived cardiorespiratory features, namely measures relating to variability and multi-scale entropy were robust and reliable features that predicted onset up to 36 hours before the clinical definition is met. The inclusion of structured data from the medical record, namely oxygenation patterns, complete blood counts, and basic metabolics further improved model performance. The combined model with 6-hour prediction horizon achieved an area under the receiver operating characteristic of 0.79 as opposed to the first 24-hour Lung Injury Prediction Score of 0.72.

## I. Introduction

Acute Respiratory Distress Syndrome (ARDS) is a life-threatening condition in which acute inflammation results in increased pulmonary vascular permeability causing diffuse alveolar damage [1]. ARDS affects 120,000 hospitalized patients annually and is an under-recognized condition, estimated to be prevalent in 10% of all patients admitted to the intensive care unit (ICU) [2]. ARDS is associated with a high mortality rate up to 45%, yet treatment options are limited for this condition, leading to a significant interest in its prediction and prevention.

The major barrier to the development of therapeutics and management strategies to prevent ARDS is the timely identification of patients who are at-risk of developing ARDS [3]. It has been reported that ARDS is often delayed, with 40% of cases entirely missed, 21% in the severe category [4]. This delay or absence of diagnosis can lead to inadvertent, unnecessary harm as well as prevent improved management such as fluid restrictive therapy and low tidal volume. Quantitative indicators that predict the disease prior to clinician suspension would allow for not only improved management, but. also, development of higher risk therapies and prognosis.

The Lung Injury Prediction Score (LIPS) has been tested for prediction of ARDS in a multi-center study with the performance measured with an area under receiver operating characteristic (AUROC) of 0.70 [5]. Machine learning algorithms have also used the Electronic Medical Record (EMR) for prediction of ARDS [6] and other diseases such as sepsis [7]. However, the EMR does have limitations due to its’ sparse nature as well as its’ indicator and behavioral biases that are involved in the ordering of labs and other clinical measures [8].

The derivation of ‘physiomarkers’, electrophysiological and hemodynamic markers, generated from continuous bedside monitoring data have been used in a number of applications, such as cardiovascular sufficiency [9], pulmonary fluid status [10] and hypotension [11]. The utility of these markers is driven by their pervasive availability, particularly those derived from electrocardiogram (ECG) and photoplethysmography [12]. However, physiomarkers have not been explored within the context of ARDS. A, and may serve as rapid indicators of risk early in the ICU encounter.

Accurate characterization of ARDS cases from a retrospective dataset in an automated fashion is a major unaddressed challenge [13]. The onset of hypoxic respiratory failure is a key clinical component in identifying patients at risk for the development of ARDS. In order to ensure accurate class labels of ARDS, clinical adjudication, that encompasses radiological images, is needed. A prediction algorithm for Hypoxic Respiratory Failure could identify candidates for clinical screening of radiological images for ARDS. The use of EMR alone can determine Hypoxic Respiratory Failure, enabling an algorithm to be developed that is economical and can be widely applicable.

In this study, novel physiomarkers, within a machine learning algorithm, were sought for prediction of the onset of Hypoxic Respiratory Failure among acute and critically ill patients. Utilizing only a single lead of the commonly available ECG waveform, we derive several candidate markers that are evaluated as predictors of ARDS at 6 and 12 hrs. prior to onset. We report the findings of that analysis, along with a data-fusion model that integrates routine clinical data abstracted from the EMR.

## II. Materials and Methods

### A. Description of Datasets

This is a retrospective study using high fidelity waveform measurements and the EMR from patients admitted to the Emory University Healthcare ICUs. The study was approved by the Emory Institutional Review Board (IRB) as non-human subjects research. All procedures were in accordance with the Helsinki Declaration of 1975.

### B. Selection Criteria

We evaluate based on a pragmatic and consensus approach that integrates data from the EMR, International Classification of Diseases (ICD) codes, lab results, and ventilator settings. An ICD 9th Edition (ICD9) code of 518.52 or ICD 10th Edition (ICD10) code of J80 was used as a candidate selection. A PaO_2_ to FiO_2_ (P/F) ratio of < 300 mmHg or a SpO_2_ to FiO_2_ (S/F) ratio of <315, and a Positive End-Expiratory Pressure (PEEP) > 5 cm H2O, where the lab value collection time and ventilator recording times occurred within 1 hour, were used as a secondary selection. The first time in which these secondary selection conditions are all met, within 1 hour, was considered the onset time of Hypoxic Respiratory Failure resulting in ARDS. Clinician adjudication was performed on a subset to determine agreement with the Berlin Definition of ARDS, through the evaluation of radiographic evidence of bilateral infiltrates, in addition to P/F < 300 and PEEP > 5.

### C. Data Processing

For case patients, waveform data available in the time window from 60 hours before onset until 12 hours after onset was included in the analysis. Due to the lack of an event within the controls, a random time was generated during the encounter, to match similar onset times within the case population. If a patient did not have at least 8 hours of usable waveform data, they were excluded from the study.

Consecutive 8-hour windows, with a stride of 1 hour, were used for classifications within the model. Statistics were reported for each feature from the 8 hourly values in the window of the case or control. For continuous features, the statistics were mean, median, minimum, maximum, skewness, variance, and kurtosis. For procedures an indicator variable was generated, e.g. admission of a vasopressor, etc. Classification of 8-hour windows was a control unless there was an overlap post prediction horizon for case patients as done in a previous study [6] and described in Supplementary Figure 1.

A full list of the EMR data is represented in Supplemental Table 1. Waveform data was obtained by processing the raw ECG captured from bedside monitors in Emory ICU’s. This raw ECG was sampled at 240 Hz. Non-overlapping windows of 5 minutes were selected for feature generation. RR intervals from the ECG were generated using the PhysioNet Cardiovascular Toolbox [14].

To identify the optimal model for the input data, a number of learning algorithms were evaluated, including Logistic Regression, Random Forest, and Xtreme Gradient Boosting [15]. Hyperparameters were tuned to optimize learning performance and described in the Supplemental Document. Feature importance and interpretability were characterized using the SHapley Additive exPlanations (SHAP) explainer [16]. Model performance was characterized using a number of algorithmic benchmarks including AUROC, Sensitivity, Specificity and Positive Predictive Value (PPV).

## III. Results

### A. Patient and Clinical Characteristics

A total of 3,708 patients with matched retrospective waveform and EMR data from October 2016 through September 2018, were screened for inclusion in the study. After applying the inclusion criteria, a total of 2,078 patients were ultimately included as shown in Figure 1.

**Figure 1.**
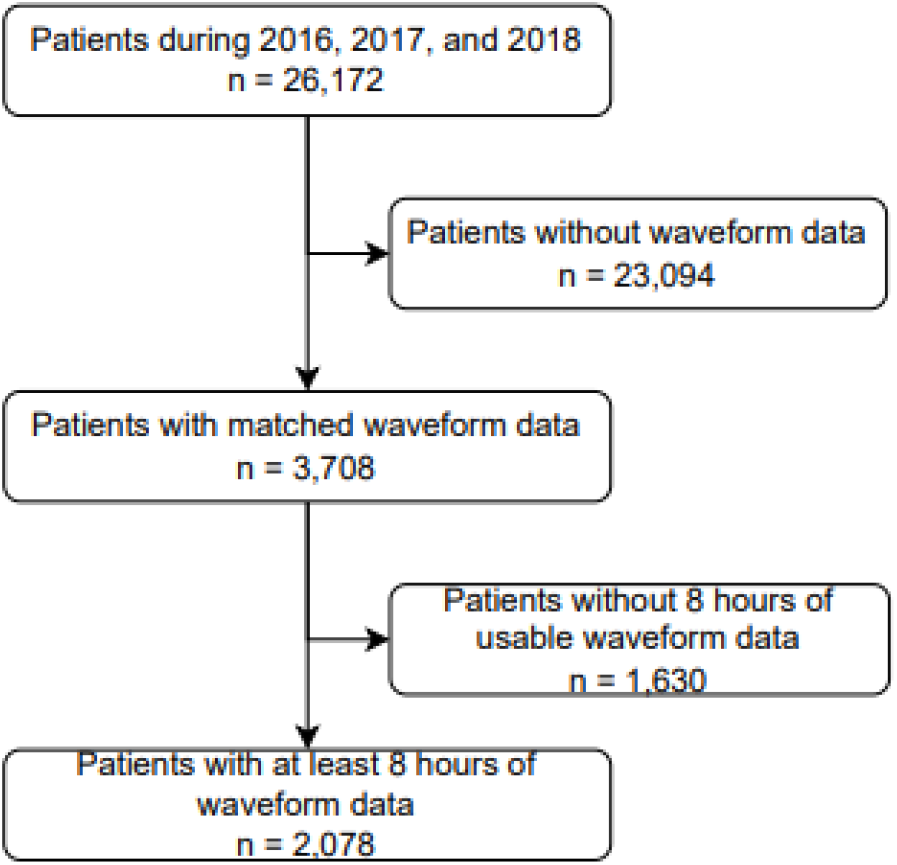
Consort Diagram

Among the final analytic cohort of 2078 patients, 128 (6.2%) were determined to have the condition based on modified Berlin criteria. Table 1 compares the clinical characteristics of the cohort stratified by the presence of ARDS compared to the absence of the condition. There were no significant differences in age and self-identified race ethnic group between those with and without the condition. Compared to those without, patients with had a higher LIPS (Mean [IQR], 6.92 [5.50 8.50] vs 5.03 [4.00 6.50], p<0.001) and lower P/F ratio (192.0 [124.7 257.3] vs 280.0 [273.3 280.0], p<0.001) consistent with worse oxygenation and possible lung injury. While in-hospital mortality was numerically higher among those with the condition compared to those without (13 (10.2%) vs. 124 (6.4%), p>0.05), this difference was not significant.

**Table 1:** Clinical characteristics of the patient cohort

|  | Total | ARDS | Control | p Value |
| --- | --- | --- | --- | --- |
| Patients (n) | 2078 | 128 | 1950 | NA |
| Males, n (%) | 1071 (51.2%) | 72 (56.3%) | 999 (51.2%) | >0.05 |
| Age, mean [IQR] | 58.4 [48.0 70.0] | 56.6 [48.8 69.0] | 58.5 [48.0 70.0] | >0.05 |
| Mechanical Ventilation, n (%) | 874 (42.1%) | 128 (100.0%) | 746 (38.3%) | <0.001 |
| In-hospital deaths | 137 (6.6%) | 13 (10.2%) | 124 (6.4%) | >0.05 |
| LIPS, mean [IQR] | 5.67 [4.00 7.00] | 6.92 [5.50 8.50] | 5.03 [4.00 6.50] | <0.001 |
| PEEP, median [IQR] | 6.0 [6.0 8.0] | 6.0 [6.0 10.0] | 6.0 [6.0 8.0] | >0.05 |
| P/F Ratio, median [IQR] | 280.0 [222.9 280.0] | 192.0 [124.7 257.3] | 280.0 [273.3 280.0] | <0.001 |
| SF Ratio, median [IQR] | 186.0 [133.3 233.8] | 139.2 [98.4 180.5] | 190.0 [140.3 240.0] | <0.001 |
| White Blood Cell count, | 9.0 [6.9 12.9] | 9.9 [7.6 16.15] | 9.0 [6.8 12.4] | <0.01 |
| Age Group, n (%): |  |  |  |  |
| 18 to 40 years | 326 (15.7%) | 23 (18.0%) | 303 (15.5%) | >0.05 |
| 41 to 60 years | 723 (34.8%) | 45 (35.2%) | 678 (34.8%) | >0.05 |
| 61 to 80 years | 876 (42.2%) | 58 (45.3%) | 818 (41.9%) | >0.05 |
| >80 years | 152 (7.3%) | 2 (1.6%) | 150 (7.7%) | >0.05 |
| Race: |  |  |  |  |
| African American | 777 (37.4%) | 43 (33.6%) | 734 (37.6%) | >0.05 |
| Caucasian | 1107 (53.3%) | 72 (56.3%) | 1035 (53.1%) | >0.05 |
| Asian | 53 (2.5%) | 4 (3.1%) | 49 (2.5%) | >0.05 |
| Other or Unknown | 141 (6.8%) | 9 (7.0%) | 132 (6.8%) | >0.05 |

Figure 2 displays the receiver operating characteristic curve for the models with prediction horizon of 12 hour before onset as well as the curve representing no prediction skill in a model. The area under the receiver operating characteristic curve is 0.59 for the model with waveform features alone and 0.76 for the combined model. The sensitivity and specificity at an operating point is also depicted in each non-trivial model, in addition to being reported in Table 2 below. On the right half of Figure 2 is a plot of AUC for models versus prediction horizon time as it is varied from 36 hours to 6 hours prior to onset. Each model was executed 30 times, the mean and confidence interval are depicted. The models are grouped together based on included features. The waveform only model results improved from the change from 36 hours to 30 hours but remained relatively constant from 30 hours to 12 hours. The 6-hour prediction horizon produced the best results and the highest change between models. In contrast to the waveform only data, the addition of EMR features resulted in increasingly improved results from 36 hours to 6 hours.

**Table 2.**
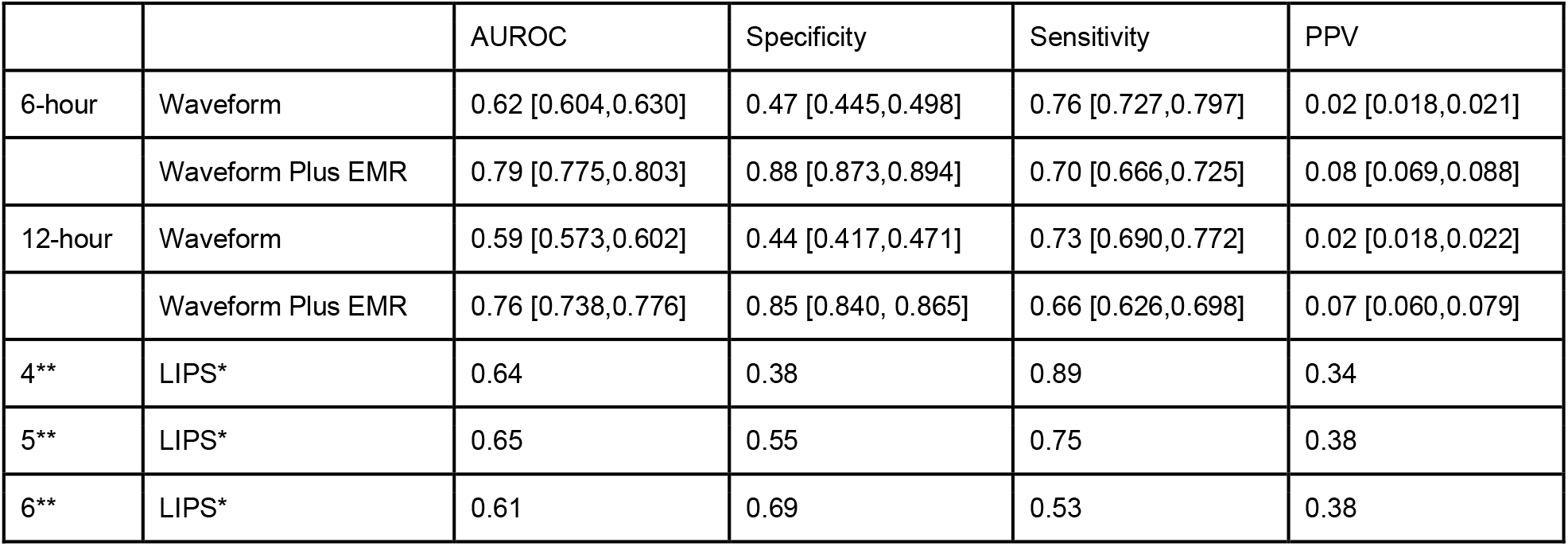
Results of 12 Hour and 6 Hour Models with LIPS Comparison *LIPS was computed using the worst features during the first 24 hours after admission. **Threshold.

**Figure 2.**
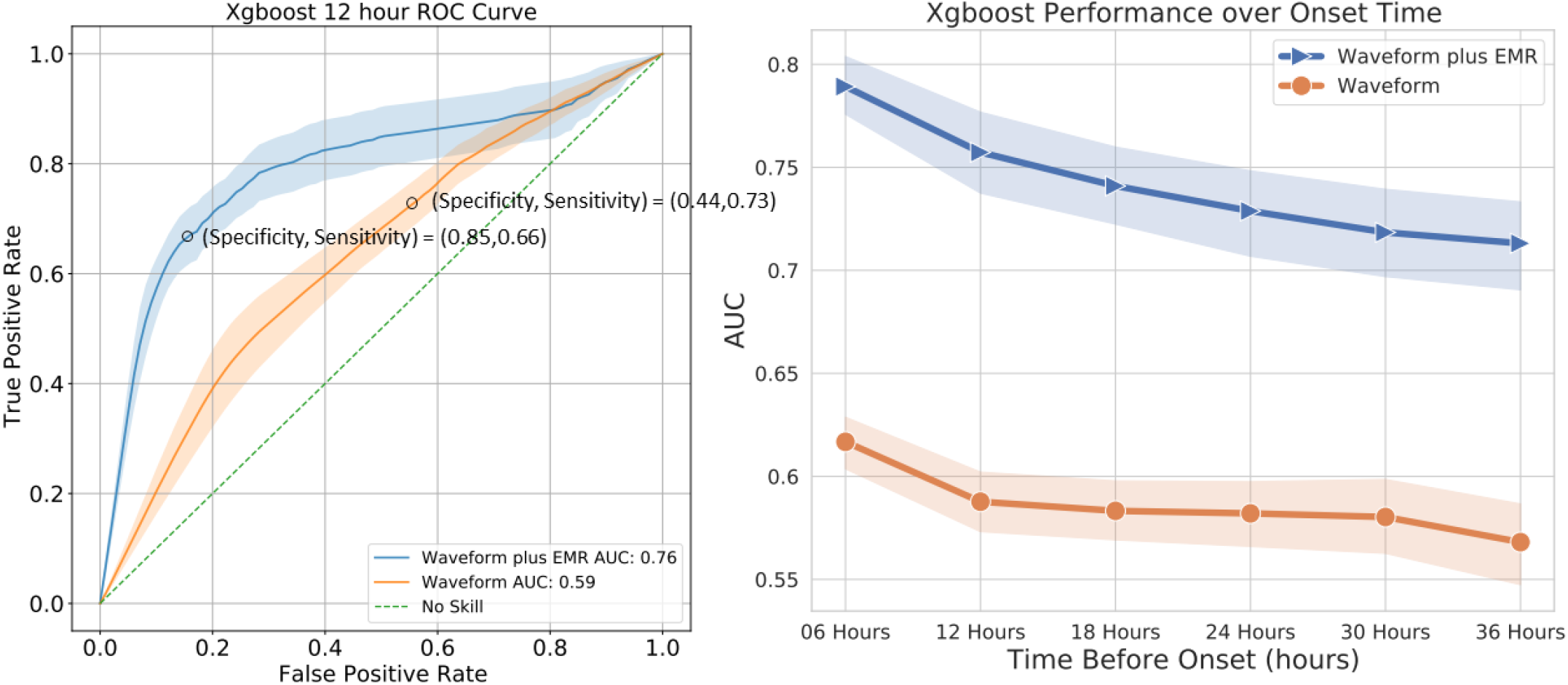
Receiver Operating Curve for 12 hour Model (left) and Trajectory of Models as Prediction Horizon Approaches Onset.

The total increase of the EMR included model was higher than the waveform only model. However, the increase from 12 hours to 6 hours was comparable for the two models.

The performance of four of the models is represented in Table 2. Models in which the prediction horizon is 12 and 6 hours prior to onset are shown. For each respective horizon, there are models including waveform features alone and those including EMR features. Considering the 12-hour prediction horizon models, the AUROC improved from 0.59, with the addition of EMR features, to 0.76. Specificity nearly doubled with the addition of EMR features, achieving better performance well outside the confidence intervals. However, waveform features alone achieved a superior sensitivity of 0.73 to 0.66, while the confidence intervals did overlap. PPV, with EMR features, was over three times without the EMR features.

Moving from the 12-hour prediction horizon to the 6 hour prediction horizon, every, respective, measure was improved. The AUC increased by 0.03 to 0.62 and 0.79 for the waveform features alone and waveform and EMR features together, respectively. Specificity also increased by 0.3 for both prediction horizons. Sensitivity increased by 0.3 to 0.76 for the model with waveform features alone while the PPV remained unchanged. The sensitivity and PPV increased by 0.1 more for the model that included EMR features when compared to without as the prediction horizon decreased.

LIPS was evaluated at three different thresholds. The AUROC and specificity performed worse for this score when compared to the combined models. The LIPS did provide a high number of true positives resulting in comparatively large PPV value and competitive sensitivity values.

Figure 3 has two representations of the SHAP values for the models with a 12-hour prediction horizon. In the waveform-only model, 4 of the top 9 features on the model are entropy measures, 3 are frequency domain measures, there is a non-linear measure and a time domain feature. The beeswarm plot depicts high entropy, of certain scales, in the signal having an impact on the model towards the negative assertion. In the combined features model, the presence of mechanical ventilation is shown to surpass all other features. There are a total of 5 features, in the top 9, that are waveform derived in this model. 3 entropy measures were in the top 9, 2 of which were also top measures in the model with waveform-only features and affected the model with the same inverse relationship. 3 EMR features of the top 9 are directly affected by mechanical ventilation: presence of the ventilator, total time on ventilation, and tidal volume. The presence of ventilation and high maximum tidal volume had a direct relationship with model output.

**Figure 3.**
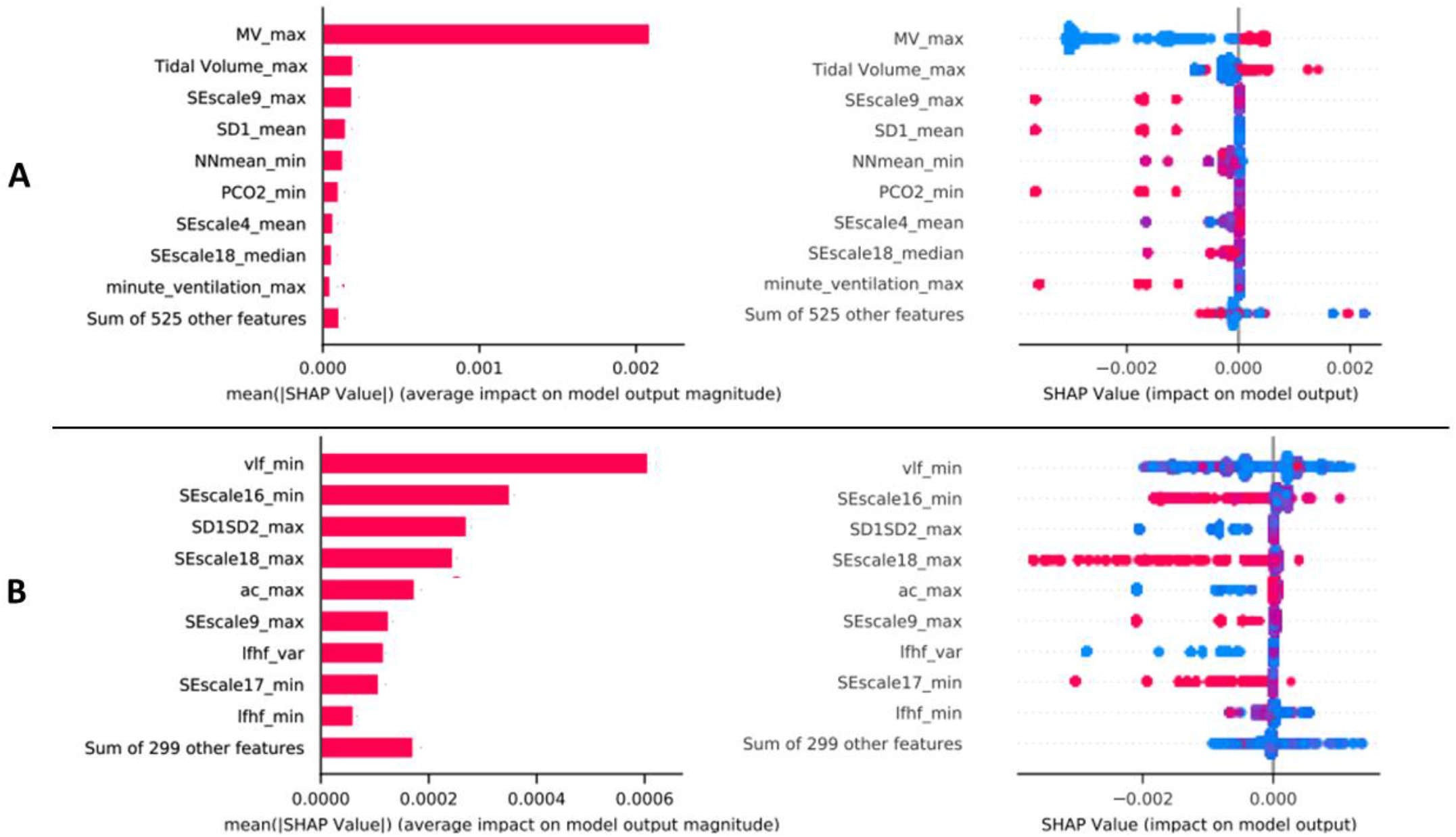
SHAP Magnitude and Beeswarm Plot for Combined Features (A) and Waveform-only Features (B) Models

Example model probabilities during the patient encounter are represented in Figure 4 for a patient who met ARDS and one who didn’t, respectively. Along with the probabilities, the relative SHAP values for top features are depicted. The probability reached a threshold value approximately 10-hour prior to when criteria was met. PaO2 variance appears to be the most influential feature in the model, although the top four features have distinct changes in the SHAP value as the probability increases. The top feature that corresponds to a waveform derived feature is a frequency domain feature, and it quantifies the power in the low frequency range of the signal. The SHAP value of this waveform derived feature is high before the probability threshold has been reached. The probability never reaches the threshold value for the patient that didn’t meet ARDS criteria.

**Figure 4.**
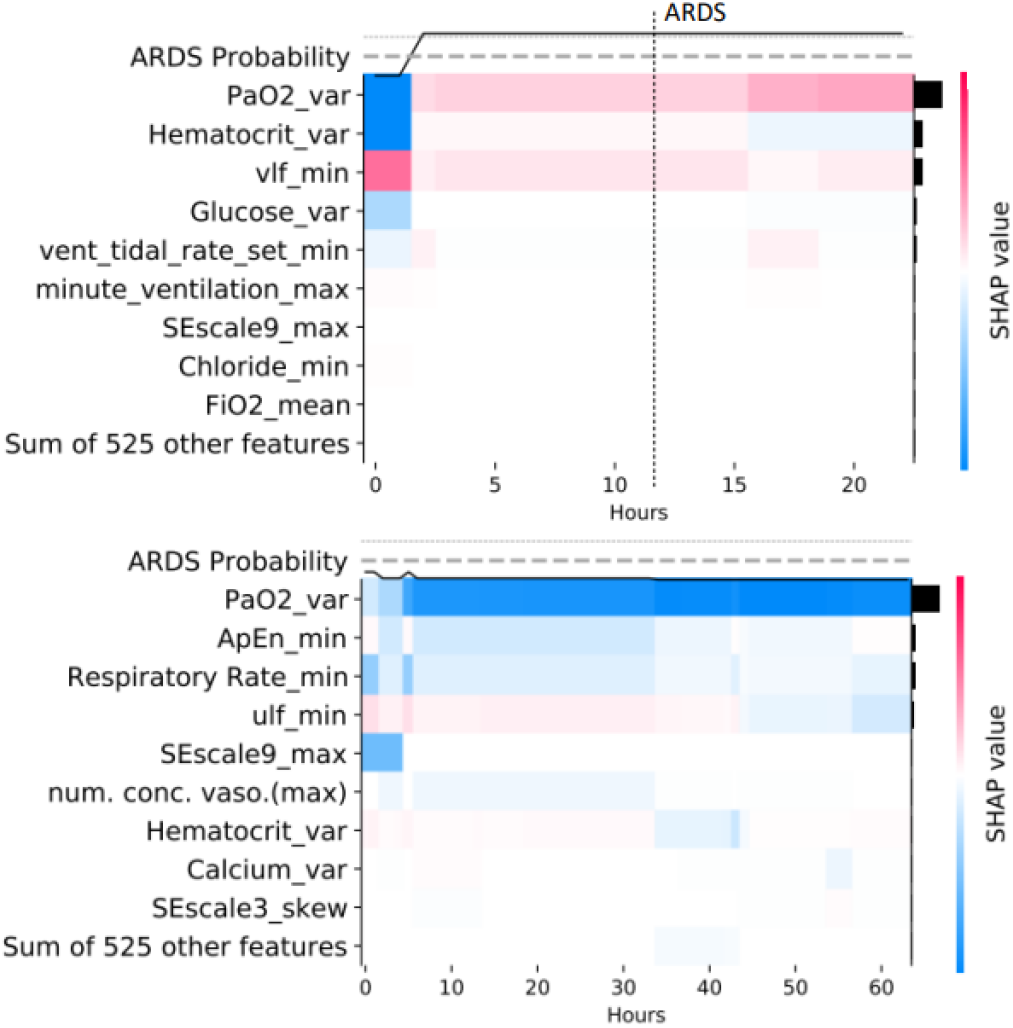
Case and Control Patient Model Probability Trajectories and Associated SHAP Values

## IV. Discussion

### A. Findings

Previous models predicting ARDS while incorporating solely electronic medical record data have shown promise [6]. However, this manuscript is the first to incorporate real-time continuous waveform data to predict the development of the disease. Continuous waveform data is not static and varies over time which is more consistent with the clinical course of patients receiving attention in acute care hospitals. The contrast with prior prediction models is significant as the data is limited in that it is captured from a single or relatively small series of discrete instances, limiting its clinical utility in predicting real-time deterioration. Incorporating continuous waveform data is novel in that it is more readily applied to the bedside as telemetry is recorded on most acute care patients, which can aid the more direct, sparse data to provide a real-time prediction.

ECG as a measurement providing waveform data was selected for the first characterization of a multi-modal machine learning model for prediction due to the cardiorespiratory physiomarkers available from the routinely and non-invasively collected signal. The implications of this work are important, namely because many predictive models which utilize physiological information have often relied on respiratory components, such as using pressure and flow waveforms [17,18]. The use of ECG may be more generalizable and have earlier utility for prediction of risk, due to their pervasive availability and the ease of extracting digital data.

The results of the models of this study shown in Figure 2 indicate that waveform data has the potential to improve prediction of ARDS. In both feature grouped models, moving the prediction window closer to onset increased accuracy. In the combined model there was increasing accuracy from 36 hours to 6 hours prior to onset. This might reflect the presence of sparse data collected more often, gradually, as the condition becomes imminent. For the waveform-only models, the increase from 12 hours to 6 hours is similar to the combined models, which may stipulate that the change in waveform signal has approximately the same effect on the prediction capability as the change in electronic medical record data. The relatively unchanged result from 12 hour to 30 hours in combination with the large drop off from 30 hours to 36 hours would be expected if the physiomarkers from waveform data reflect the condition presence earlier than the sparse data of electronic medical record data and provide a period of elevated risk well before onset from which treatment can be delivered.

The top two features of the combined model, shown in Figure 3, are mechanical ventilator derived, which is consistent with the definition and treatment of the condition. The large magnitude of mechanical ventilator presence SHAP value is driving the specificity and true negative rate at the expense of the sensitivity. Waveform derived features compose 5 of the top 9 features, and these features have more of an effect on the model than features directly related to the criteria of the condition such as the PaO2 to FiO2 ratio.

This study demonstrates the applicability of clinical waveform data in predicting ARDS. Models encompassing only EEG derived features increase predictive ability as the onset of ARDS is approached. A similar approach with waveform data for prediction of sepsis has been studied [19]. The sepsis study included the invasive arterial blood pressure waveform in addition to EEG, enabling the calculation of pulse travel time. The use of invasive waveform data will limit the patient population with which the prediction algorithm could be applied. However, a substitute non-invasive waveform signal may allow for similar pulse travel time features to be used. This highlights the need for future work to elucidate the best features for the study of the development of ARDS. In addition, the physiology behind the feature will enable better understanding of any prediction algorithm.

### B. Limitations

Selection of case patients in this study was chosen by application of a pragmatic and consensus-based definition utilizing a combination of clinical and ICD data. This design allows the algorithm to be widely applicable across different hospital systems and for large sample sizes. This study is aimed at predicting ARDS; however, we acknowledge that clinical data utilizing the PF ratio and PEEP, and conformal verification through ICD code may underselect for patients who developed mild cases of ARDS. In order to mitigate this concern, we evaluated clinical adjudication on a subset of the data and found acceptable concordance with 80% agreement.

A fixed time frame, with a maximum of 72 hours, was chosen for consideration in this study due to the average population-level incident of ARDS being recorded within 2.5 days of ICU admission. However, Hypoxic Respiratory Failure may develop later in an ICU visit, and future work will incorporate that into the probability from initial ICU admittance, when waveform data is typically first available, for increased applicability.

## Supporting information

Supplemental Document (Data Processing)

## Data Availability

Access to de-identified Emory University cohort may be made available via approval from Emory Institutional Review Board (IRB) and Data Oversight Committee (DOC). Access to the computer code used in this research is available upon request to the corresponding author.

## Notes

This study was submitted for review on November 5, 2022. R Kamaleswaran was supported by the National Institutes of Health under Award Numbers R01GM139967 and UL1TR002378. R Kamaleswaran and C. E. Marshal were supported by Surgical Critical Care Initiative, funded through the Department of Defense’s Health Program—Joint Program Committee 6/Combat Casualty Care (USUHS HT9404-13-1-0032 and HU0001-15-2-0001).

### Competing Interest Statement

The authors have declared no competing interest.

### Funding Statement

R. Kamaleswaran was supported by the National Institutes of Health under Award Numbers R01GM139967 and UL1TR002378. R Kamaleswaran and C. E. Marshal were supported by Surgical Critical Care Initiative, funded through the Department of Defense Health Program Joint Program Committee 6/Combat Casualty Care (USUHS HT9404-13-1-0032 and HU0001-15-2-0001).

### Author Declarations

The study was approved by the Emory Institutional Review Board (IRB) as non-human subjects research.

