## Supplemental Document (Data Processing) for "A Machine Learning Algorithm to Predict Hypoxic Respiratory Failure and risk of Acute Respiratory Distress Syndrome (ARDS) by Utilizing Features Derived from Electrocardiogram (ECG) and Routinely Clinical Data"

EMR data included vital signs, lab results, ventilator status and settings, demographics, and existing conditions. A full list of the EMR variables is represented in Supplemental Table 1. For data such as lab results, where there could be a lag time between the collection time and when the results are reported, collection time was used. Preprocessing was performed to remove outliers (most likely due to input errors). Acceptable ranges (with consideration of abnormal condition) for most vitals and lab variables were derived from clinical input. Outing values from these acceptable ranges were removed from the dataset. All data was initially aggregated on an hourly basis. If multiple measurements were taken within an hour for a particular variable, the median value was taken. Statistics were reported for each feature from the 8 hourly values in the window of the case or control. For continuous features, the statistics were mean, median, minimum, maximum, skewness, variance, and kurtosis. This process is depicted in Supplemental Figure 1. For procedures an indicator variable was generated, e.g. admission of a vasopressor, etc.

All waveform derived variables were continuous and were treated in the same fashion as the EMR continuous variables to create features. A total list of variables is available in Supplemental Figure 2. Many of the variables can be separated into categories such as time domain, frequency domain, or entropy based.

**Supplemental Table1: EMR Variables**

| Category | Sub-category | Variable |
|---|---|---|
| 6 statistics | Vitals | BMI<br>Daily Weight<br>Heart Rate<br>Mean Arterial Pressure<br>Respiratory Rate<br>Systolic Blood Pressure<br>Temperature |
| | Lab Values | Albumin<br>Bicarbonate<br>Bilirubin<br>Calcium<br>Chloride<br>Creatinine<br>FIO2<br>Glucose<br>Hematocrit<br>Lactate<br>PCO2<br>PaO2<br>Platelet<br>Potassium<br>Sodium<br>White Blood Count |
| | Ventilator Settings | FIO2<br>GCS<br>PF Ratio<br>Vent Rate<br>Vent Tidal Rate<br>Tidal Volume |
| | Metric | GCS |
| Maximum value | Concurrent Conditions | Cardiac Arrhythmia |

| | | |
|---|---|---|
| | | Chronic Pulmonary<br>Congestive Heart Failure<br>Diabetes complicated<br>Diabetes Uncomplicated<br>Hypertension<br>Liver Disease<br>Metastatic Cancer<br>Obesity<br>Renal Failure<br>Rheumatoid Arthritis<br>Valvular Disease |
| | Treatment | Dobutamine<br>Dopamine<br>Epinephrine<br>Midodrine<br>Milrinone<br>Norepinephrine<br>Number of Concurrent Inotropes<br>Number of Concurrent Vasopressors<br>Phenylephrine<br>Vasopressin<br>Ventilation Duration (Min.) |
| Demographics | Demographics | Age<br>Gender<br>Race |

**Supplemental Figure 1: Depiction of Hourly Resampling (A), Accounting for Missing Data (B), Windowing Data for Classification (C), and Statistics Used for Continuous Variables That Make Up Features (D)**

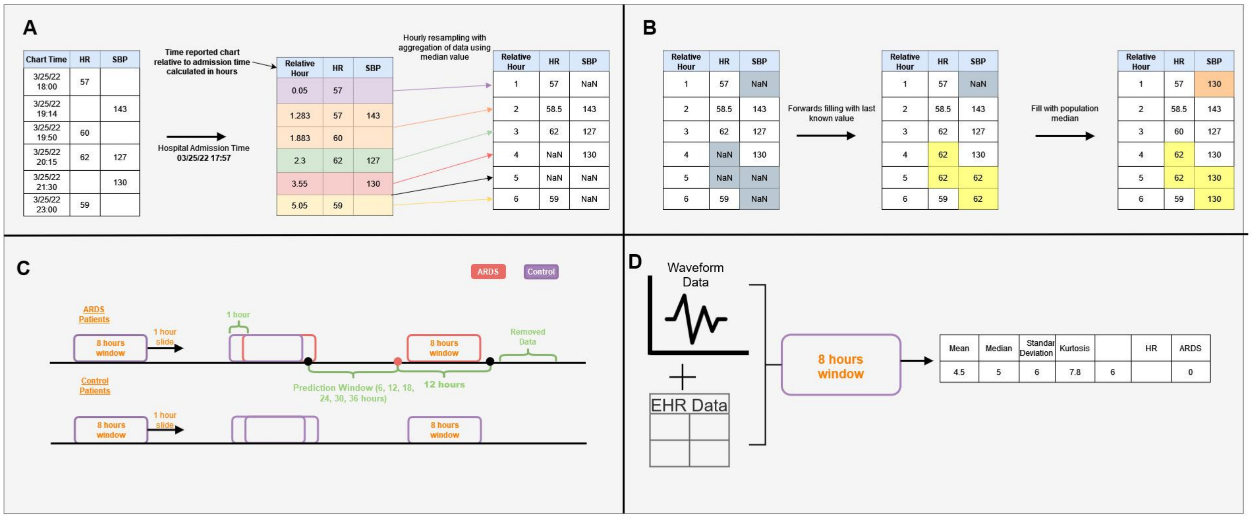

### Supplemental Table 2: Waveform Derived Variables

| Category | Variable |
|---|---|
| Time Domain | NNmean |
| | NNmedian |
| | NNmode |
| | NNvariance |
| | NNskew |
| | NNkurt |
| | NNiqr |
| | SDNN |
| | pnn50 |
| | RMSSD |
| | avgsqi |
| Frequency Domain | ulf |
| | vlf |
| | lf |
| | hf |
| | lfhf |
| | ttlpwr |
| Other | ac |
| | dc |
| | SD1 |
| | SD2 |
| | SD1SD2 |
| Entropy | SampEn |
| | ApEn |
| | SEscale1 |
| | SEscale2 |

| | |
|---|---|
| | SEscale3 |
| | SEscale4 |
| | SEscale5 |
| | SEscale6 |
| | SEscale7 |
| | SEscale8 |
| | SEscale9 |
| | SEscale10 |
| | SEscale11 |
| | SEscale12 |
| | SEscale13 |
| | SEscale14 |
| | SEscale15 |
| | SEscale16 |
| | SEscale17 |
| | SEscale18 |
| | SEscale19 |
| | SEscale20 |
